# COVID-19 booster dose antibody response in pregnant, lactating, and nonpregnant women

**DOI:** 10.1101/2022.05.17.22275154

**Authors:** Caroline Atyeo, Lydia L. Shook, Nadege Nziza, Elizabeth A. Deriso, Cordelia Muir, Arantxa Medina Baez, Rosiane S. Lima, Stepan Demidkin, Sara Brigida, Rose M. De Guzman, Madeleine D. Burns, Alejandro B. Balazs, Alessio Fasano, Lael M. Yonker, Kathryn J. Gray, Galit Alter, Andrea G. Edlow

## Abstract

**BACKGROUND:** While emerging data during the SARS-CoV-2 pandemic have demonstrated robust mRNA vaccine-induced immunogenicity across populations, including pregnant and lactating individuals, the rapid waning of vaccine-induced immunity and the emergence of variants of concern motivated the use of mRNA vaccine booster doses. Whether all populations, including pregnant and lactating individuals, will mount a comparable response to a booster dose is not known.

**OBJECTIVE:** We sought to profile the humoral immune response to a COVID-19 mRNA booster dose in a cohort of pregnant, lactating, and age-matched nonpregnant women.

**STUDY DESIGN:** We characterized the antibody response against ancestral Spike and Omicron in a cohort of 31 pregnant, 12 lactating and 20 nonpregnant age-matched controls who received a BNT162b2 or mRNA-1273 booster dose after primary COVID-19 vaccination. We also examined the vaccine-induced antibody profiles of 15 maternal:cord dyads at delivery.

**RESULTS:** Receipt of a booster dose during pregnancy resulted in increased IgG1 against Omicron Spike (post-primary vaccination vs post-booster, p = 0.03). Pregnant and lactating individuals exhibited equivalent Spike-specific total IgG1, IgM and IgA levels and neutralizing titers against Omicron compared to nonpregnant women. Subtle differences in Fc-receptor binding and antibody subclass profiles were observed in the immune response to a booster dose in pregnant compared to nonpregnant individuals. Analysis of maternal and cord antibody profiles at delivery demonstrated equivalent total Spike-specific IgG1 in maternal and cord blood, yet higher Spike-specific FcγR3a-binding antibodies in the cord relative to maternal blood (p = 0.002), consistent with preferential transfer of highly functional IgG. Spike-specific IgG1 levels in the cord were positively correlated with time elapsed since receipt of the booster dose (Spearman R 0.574, p = 0.035).

**CONCLUSIONS:** These data suggest that receipt of a booster dose during pregnancy induces a robust Spike-specific humoral immune response, including against Omicron. If boosting occurs in the third trimester, higher Spike-specific cord IgG1 levels are achieved with greater time elapsed between receipt of the booster and delivery. Receipt of a booster dose has the potential to augment maternal and neonatal immunity.

## Introduction

Pregnant individuals are particularly vulnerable to COVID-19, as they are at increased risk of severe disease as well as adverse pregnancy outcomes including stillbirth.^1–4^ Despite CDC and ACOG recommendations encouraging all individuals who are pregnant, recently pregnant, or considering pregnancy to receive a COVID-19 vaccine,^5,6^ vaccine coverage of pregnant individuals has lagged behind that of the general adult population, with 69% of pregnant individuals vaccinated as of February 2022, compared to 82% of the nonpregnant population.^7,8^ With the emergence of SARS-CoV-2 variants of concern and evidence of waning vaccine-induced immunity in the general population, mRNA vaccine boosters are now recommended for all adults, including pregnant individuals, at least 5 months after completion of the initial vaccine series.^9^ However, as of late February 2022, only 49% of fully vaccinated pregnant individuals had received a booster dose, with uptake lowest in non-Hispanic Black and Hispanic individuals.^10^

Importantly, recent data from Israel clearly indicate improved effectiveness against severe disease after the third and even a fourth booster dose in the general adult population.^11,12^ Whether pregnant and lactating individuals, who can exhibit dampened immunity to vaccines,^16^ mount a comparably protective response to the booster dose is not known. Studies of pregnant and lactating women receiving COVID-19 vaccination demonstrated robust immunogenicity to mRNA vaccines, comparable to nonpregnant women.^13,14^ However, comprehensive profiling of the immune response to primary vaccination in pregnant and lactating women revealed reduced Fc-receptor binding and subclass selection differences, suggesting that development of a fully mature immune response may be delayed in these groups.^15^

To determine whether pregnant or lactating individuals respond effectively to a COVID-19 booster dose, we comprehensively profiled the vaccine-induced antibody response against ancestral Spike and Omicron (B.1.1.529) in a cohort of 63 individuals (31 pregnant, 12 lactating and 20 nonpregnant age-matched control women) who received a BNT162b2 or mRNA-1273 booster dose. In addition, we characterized the transfer of vaccine-induced antibodies in 15 maternal:cord dyads at delivery.

## Materials and Methods

### Participant recruitment and study design

Women at two tertiary care hospitals were approached for enrollment in an Institutional Review Board (IRB)–approved (protocol #2020P003538) COVID-19 pregnancy biorepository study. Eligible women were pregnant, lactating, or nonpregnant and of reproductive age (18 to 45) and receiving a COVID-19 mRNA vaccine booster dose (August - December 2021). Eligible participants were identified by practitioners at the participating hospitals or were self-referred. Blood was collected approximately 4 weeks after the booster dose and/or at delivery. For participants who delivered during the study period (n=15), maternal and umbilical cord blood was collected at delivery.

### Antigen-specific isotype titer and FcR-binding

Antigen-specific isotype titer and FcR-binding was measured by a multiplex Luminex, as previously described.^17^ Briefly, carboxylated Magplex microspheres were covalently linked to antigen by ester-NHS linkages using sulfo-NHS (Thermo Fisher) and EDC (Thermo Fisher). Immune complexes were formed by by adding antigen-coupled microspheres and appropriately diluted plasma (1:100 for IgG2, IgG3, IgA1 and IgM; 1:500 for IgG1; 1:1000 for FcRs). 384-well plates were incubated overnight at 4°C, shaking at 700 rpm. Plates were then washed with assay buffer (1x PBS with 0.1% BSA/0.02% Tween-20). PE-coupled mouse anti-human detection antibodies were added to detect antigen-specific isotype titer (Southern Biotech). To detect antigen-specific FcR-binding, Avi-tagged FcRs (Duke Human Vaccine Institute) were biotinylated with a BirA500 kit (Avidity). The biotinylated FcRs were then fluorescently tagged using streptavidin-PE (Agilent) and added to immune complexes. Fluorescence was read using an iQue (Intellicyt). Data are reported as median fluorescence intensity (MFI). The assay was run in duplicate, and the average of the replicates are reported.

### SARS-CoV-2 Omicron pseudovirus neutralization assay

Omicron spike pseudovirus neutralization assay was performed as previously described.^18^ A pseudovirus encoding Omicron Spike was produced by transfecting 293T cells with an Omicron Spike expression plasmid, a lentiviral backbone encoding CMV-Luciferase-IRES-ZsGreen and lentiviral helper plasmids. Diluted plasma was incubated with the Omicron pseudovirus for 1 hour, followed by addition of 293T-ACE2 cells. Cells and pseudovirus were incubated at 37°C for 48 hours. Cells were lysed and luciferase expression was assessed using a Spectramax L luminometer. The NT50 values were calculated in GraphPad Prism (version 8.0).

### Statistical Analysis

Statistical analysis was performed in R (version 4.0.0) or GraphPad Prism (version 8.0). Prior to multivariate analysis, Luminex data was log10-transformed and all data was centered and scaled. For univariate analysis, significance was determined by Kruskal-Wallis or Mann-Whitney U test. For multivariate analysis, the systemseRology R package (v1.0) (https://github.com/LoosC/systemsseRology) was used. Least absolute shrinkage and selection operator (LASSO) feature selection was performed 100 times and features selected were those chosen at least 50% of the repetitions performed.

## Results

### Similar vaccine-induced Spike-specific antibodies in pregnant, lactating and non-pregnant women after booster dose

Cohort demographic characteristics and clinical information for the 31 pregnant, 12 lactating and 20 nonpregnant age-matched individuals included in the study are reported in Table 1. There were no differences in age, race, or ethnicity between groups. Although post-booster samples were collected at least 2 weeks from receipt of the booster dose, samples from nonpregnant individuals were collected approximately 10 days later than pregnant and lactating individuals. Of the 31 pregnant participants, 77% had completed primary vaccination prior to conception. Pregnant individuals delivering during the study period (n = 15) had received the booster dose between 32 and 38 weeks of pregnancy.

**Table 1.**
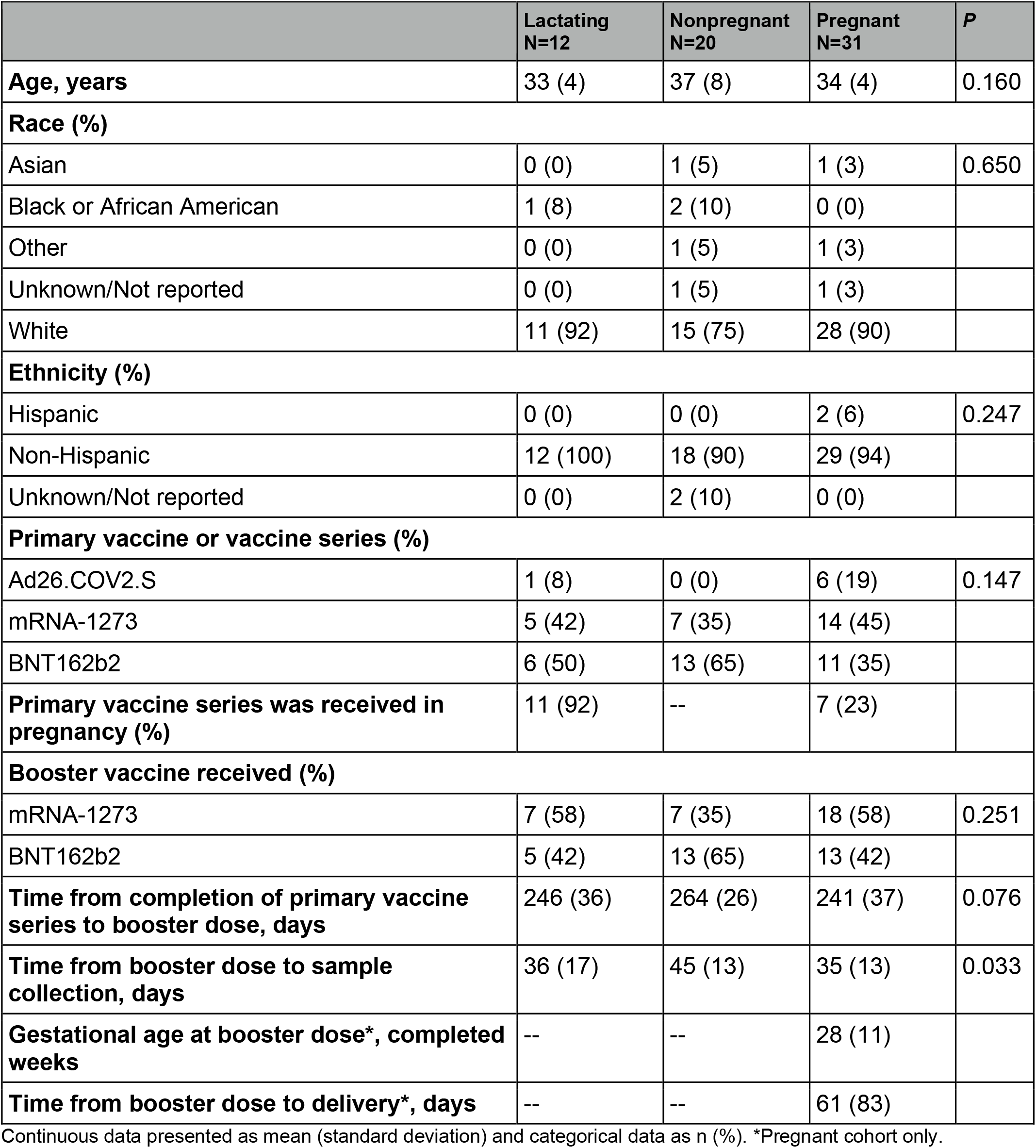
Demographic characteristics and clinical information of the study cohort.

To determine how the antibody response to a booster dose compared to the response produced after the primary vaccine series in pregnant individuals, we plotted the antibody response in a subset of 5 pregnant individuals 2 to 6 weeks after completion of primary mRNA vaccination (V2), and the response in the same individuals 4 weeks after the booster dose (V3). The booster dose induced higher IgG1and IgA levels than those induced by primary vaccination, against both the ancestral SARS-CoV-2 Spike (IgG1 V2 vs V3: p=0.06, IgA V2 vs V3: p=0.06) and the Receptor Binding Domain (RBD) (IgG1 V2 vs V3: p=0.06, IgA V2 vs V3: p=0.06), and a stable IgM response against Spike and RBD (Figure 1A, Figure S1A).

**Figure 1.**
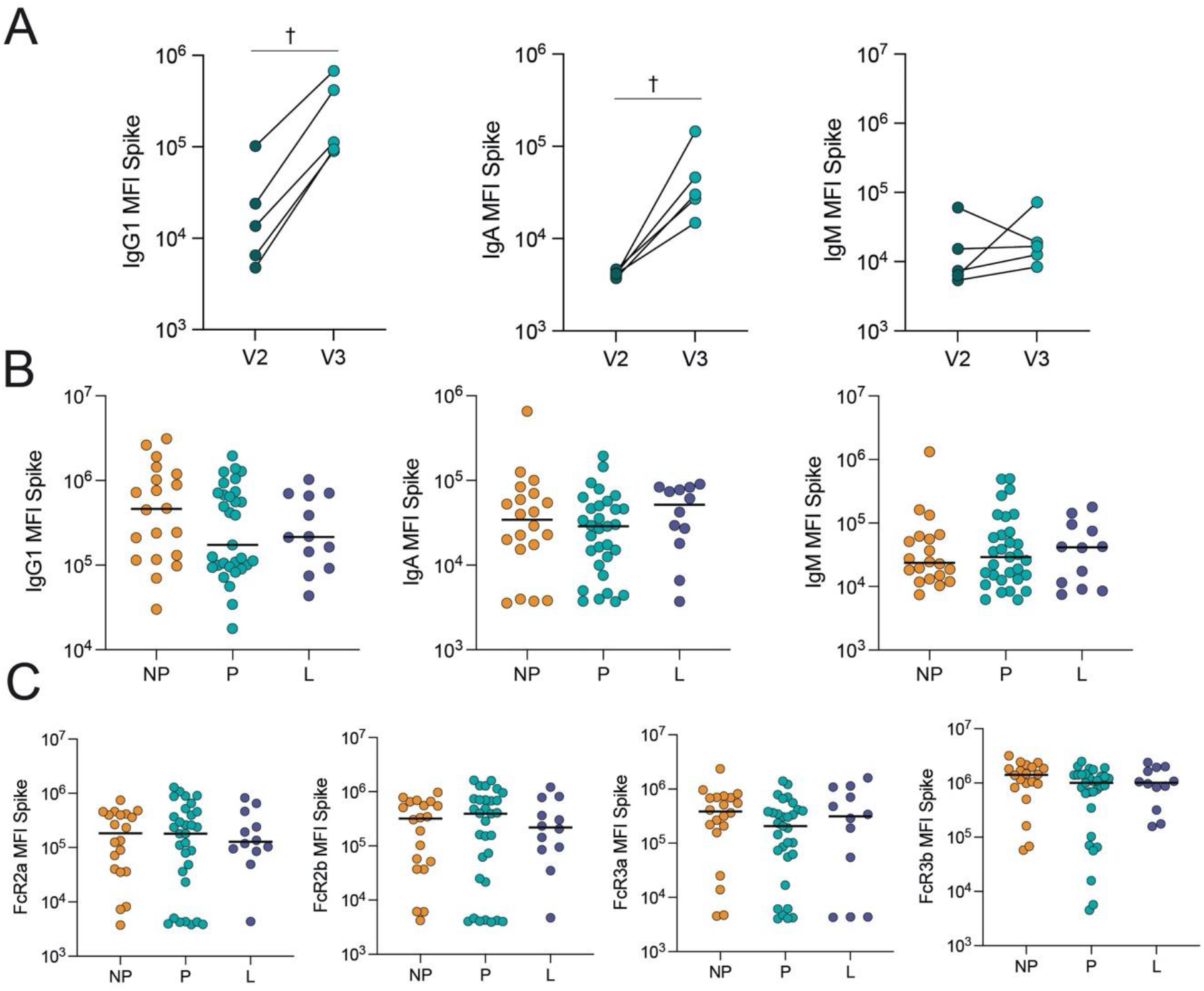
COVID-19 booster vaccine induces a similar Spike-specific antibody response in pregnant, lactating and nonpregnant individuals. A. The dot plots show the peak IgG1, IgA and IgM response against Spike in 5 pregnant individuals after receiving the second dose of a primary mRNA vaccine series (V2) and after the booster dose (V3). Lines connect samples from the same individual. Significance was determined by a Wilcoxon signed-rank test. Differences did not reach statistical significance. † p = 0.06. B. The dot plots show IgG1, IgA and IgM levels against Spike in nonpregnant (NP), pregnant (P) and lactating (L) individuals. Horizontal line represents the median for each group. Significance was determined by a Kruskal-Wallis test. No comparisons were significant. C. The dot plots show the FcR-binding of antibodies against Spike in nonpregnant (NP), pregnant (P) and lactating (L) individuals. Horizontal line represents the median for each group. Significance was determined by a Kruskal-Wallis test. No comparisons were significant.

Previous data from our group revealed that after the primary mRNA vaccine series, pregnant and lactating women induced similar IgG, IgA and IgM levels but slower evolution of Fcγ Receptor (FcγR)-binding antibodies to Spike compared to nonpregnant women.^15^ Therefore, we next aimed to determine whether a booster dose could compensate for this observed deficit in immunity observed in pregnant individuals. We observed a slightly, but not significantly, lower IgG1 against Spike (pregnant: 1.7×10^5^ MFI, nonpregnant: 4.6×10^5^ MFI, lactating: 2.2×10^5^ MFI) and RBD (pregnant: 1.3×10^5^ MFI, nonpregnant: 3.5 ×10^5^ MFI, lactating: 2.1 ×10^5^ MFI) in pregnant women compared to nonpregnant and lactating women, although similar IgA and IgM levels were observed in all three groups (Figure 1B, Figure S1B). Moreover, FcγR-binding against Spike and RBD was nearly equivalent across the groups (Figure 1C, Figure S1C).

### A booster dose induces an increase in Omicron-specific IgG1 and an equivalent Omicron-specific antibody response between pregnant and nonpregnant individuals

Emerging data point to a critical role for boosting not only in augmenting the absolute amount of antibody to ancestral Spike, but also in improving the breadth of the response to variants of concern (VOCs).^18–20^ In particular, the ability of the mRNA booster dose to provide protection against Omicron, which has become the predominant strain in the United States, is crucial. Boosting resulted in a robust increase in Omicron Spike-specific IgG1 (median IgG1 V2 vs V3: 1.4×10^4^ MFI vs 1.1×10^5^ MFI, p=0.03), but not IgA or IgM (Figure 2A). Comparison of Omicron-specific binding profiles across nonpregnant, pregnant, and lactating women pointed to slightly lower Omicron Spike-specific IgG1 in pregnant individuals (pregnant: 1.2 ×10^5^ MFI, nonpregnant: 2.0 ×10^5^ MFI, lactating: 1.5 ×10^5^ MFI) but equivalent IgA and IgM titers and neutralizing activity across the 3 groups (Figure 2B, 2C). Omicron Spike-specific FcγR-binding was equivalent between groups (Figure 2D).

**Figure 2.**
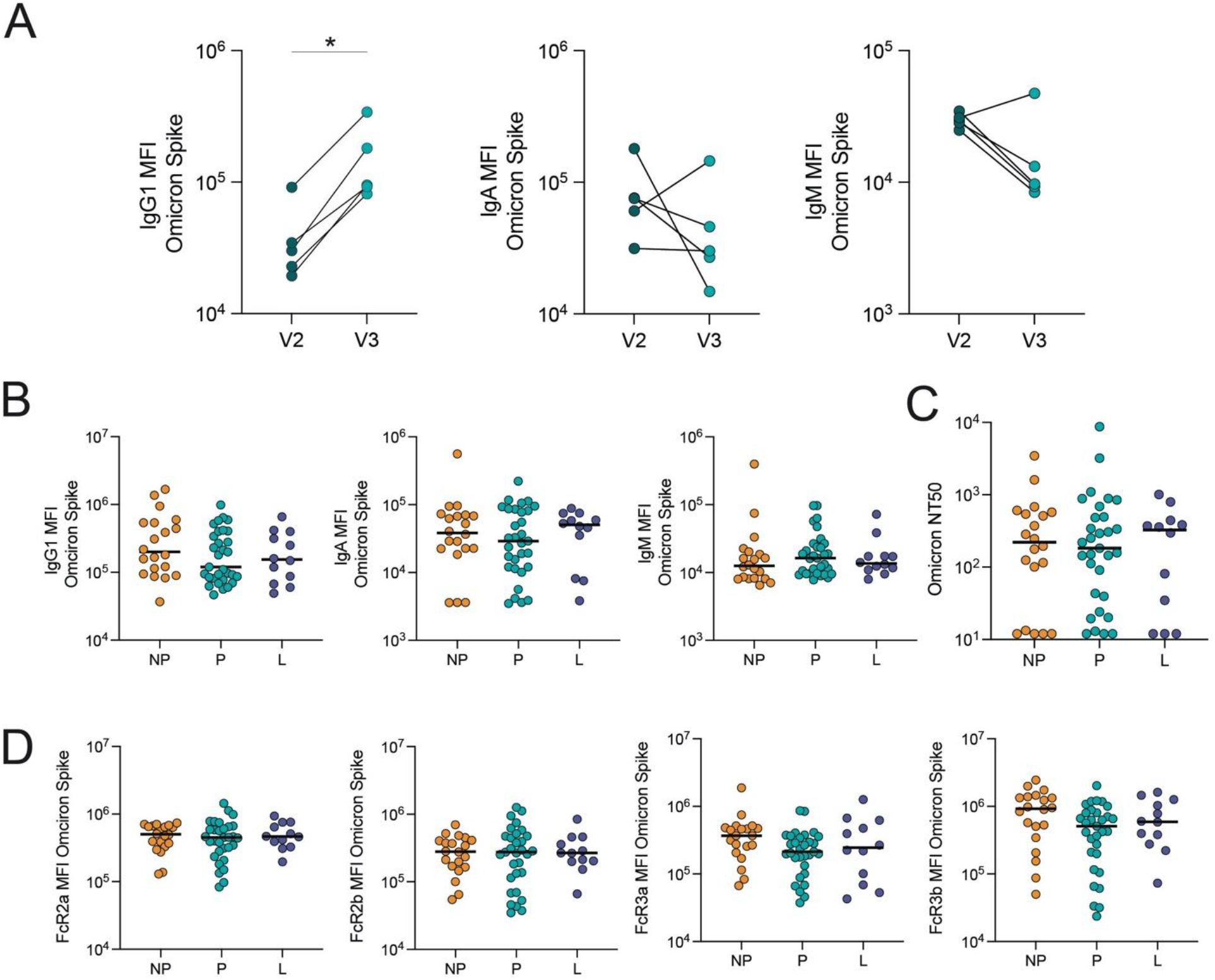
COVID-19 booster vaccine induces a similar Omicron Spike-specific antibody response in pregnant, lactating and nonpregnant individuals. A-B. The dot plots show the peak IgG1, IgA and IgM response against Omicron Spike in 5 pregnant individuals after receiving the second dose of a primary mRNA vaccine series (V2) and after the booster dose (V3). Lines connect samples from the same individual. Significance was determined by a Wilcoxon signed-rank test, * p<0.05. B. The dot plots show IgG1, IgA and IgM levels against Omicron Spike in nonpregnant (NP), pregnant (P) and lactating (L) individuals. Horizontal line represents the median for each group.Significance was determined by a Kruskal-Wallis test. No comparisons were significant. C. The dot plots show the NT50 against an Omicron Spike pseudovirus in nonpregnant (NP), pregnant (P) and lactating (L) individuals. Horizontal line represents the median for each group. Significance was determined by a Kruskal-Wallis test. No comparisons were significant. D. The dot plots show the FcR-binding of antibodies against Omicron Spike in nonpregnant (NP), pregnant (P) and lactating (L) individuals. Horizontal line represents the median for each group. Significance was determined by a Kruskal-Wallis test. No comparisons were significant.

### Differences in antibody class-switching are observed in pregnant individuals

The observation of subtle univariate differences in vaccine-induced IgG1 responses after boosting (Figure 1) prompted the further dissection of differences by multivariate modeling. First, to understand if certain antibody levels were different between the three groups, we used LASSO to define the minimal set of antibody features that were the most different between groups. From this analysis, LASSO selected 10 of the total 75 antibody features per sample that separated the antibody profiles in each of the three groups (Figure 3A). We then performed univariate analysis for each of the LASSO-selected features (Figure 3B). From this analysis, we identified that there was an elevation of FcγR-binding/IgG1 against Omicron RBD and FcγR-binding/IgG2 ancestral strain N-terminal domain (NTD) in nonpregnant compared to pregnant individuals. Moreover, we observed a shift towards an elevation of IgG3 in lactating/pregnant individuals and a slight elevation of IgM in pregnant individuals. Taken together, these data may suggest more robust vaccine-induced selection of pre-existing memory B cells and antibody class switching in nonpregnant individuals, versus enhanced selection of naïve B cell responses in pregnant individuals (see Comment). The significant increase in IgG2 S2 in lactating individuals compared to pregnant individuals suggests a return towards the nonpregnant immune state during lactation.

**Figure 3.**
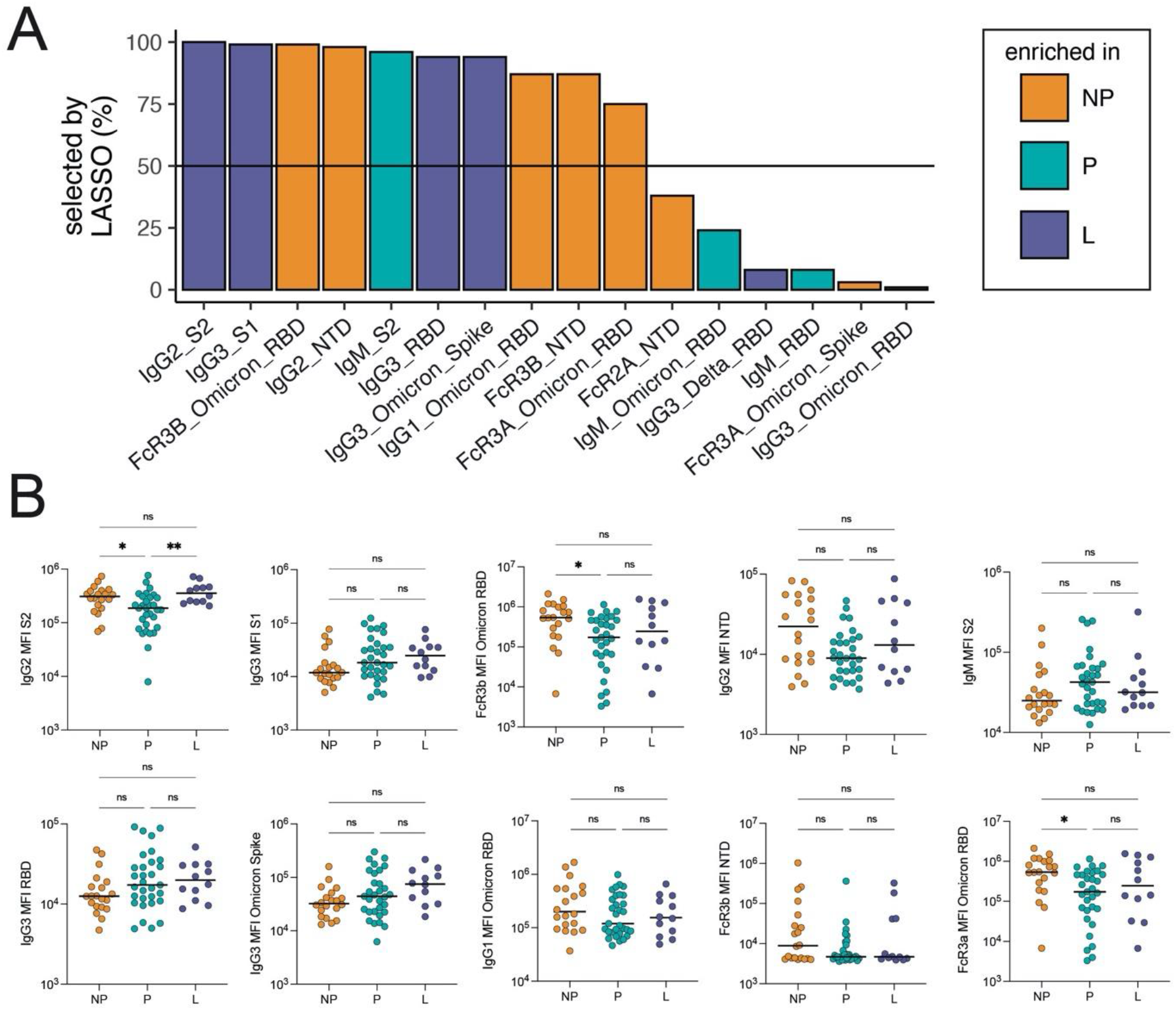
Differences in the antibody response in nonpregnant and pregnant individuals after boosting. A. Least absolute shrinkage and selection operator (LASSO) was used to select the antibody features that separated the three groups: nonpregnant (NP), pregnant (P) and lactating (L) individuals. LASSO was performed 100 times, and the barplot shows the percentage that each feature was selected (for features selected at least once). The horizontal line represents the 50% cutoff used to define the top features. The color of the bar represents the group in which the feature is the most elevated. B. The dot plots show the univariate analysis of the LASSO-selected features (A). Horizontal line represents the median for each group. Horizontal line represents the median for each group. Significance was determined by a Kruskal-Wallis test. P-values were corrected for multiple testing by the Benjamini-Hochberg method, * p<0.05, ** p<0.01.

### Boosting results in transfer of antibodies to the cord in a time-dependent manner

IgG1 against ancestral Spike in matched maternal and cord plasma obtained at delivery from 15 pregnant individuals who delivered during the study period were equivalent (Figure 4A). Similarly, we observed an equivalent level of neutralizing antibodies against Omicron in both the maternal and cord blood (Figure 4B). IgA and IgM against ancestral Spike were not transferred to the cord, as expected (Figure S2A). Analysis of FcγR-binding antibodies revealed that although levels of FcγR2a-, FcγR2b- and FcγR3b-binding antibodies against Spike were equivalent in maternal and cord blood, a higher concentration of FcγR3a-binding antibodies was observed in cord relative to maternal blood (cord: 4.9×10^5^ MFI, maternal: 1.0×10^5^ MFI, p = 0.002), consistent with efficient transplacental transfer of these antibodies (Figure 4C). We observed similar transfer patterns of antibodies against Omicron Spike (Figure S2B/C).

**Figure 4.**
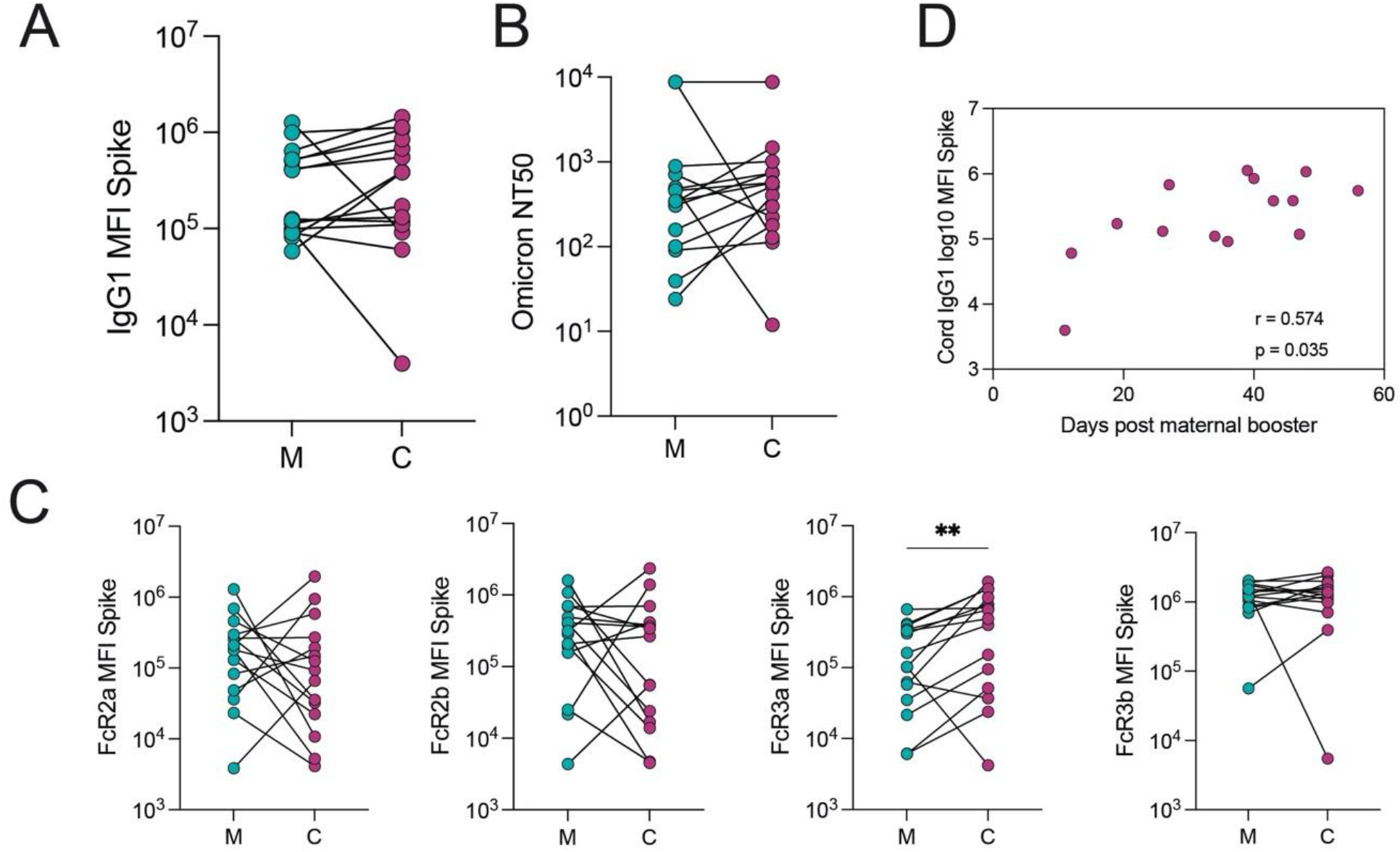
The transfer of Spike-specific antibodies to the cord after vaccination in the third trimester. A. The dot plots show the IgG1 against Spike in maternal (M) and cord blood (C). Lines connect matched maternal:cord dyads (n=15). Significance was determined by a Wilcoxon signed-rank test. B. The dot plots show the NT50 against an Omicron Spike pseudovirus in maternal (M) and cord blood (C). Lines connect matched maternal:cord dyads (n=15). Significance was determined by a Wilcoxon signed-rank test. C. The dot plots show the FcR-binding titer against Spike in maternal (M) and cord blood (C). Lines connect matched maternal:cord dyads (n=15). Significance was determined by a Wilcoxon signed-rank test followed by a Benjamini-Hochberg correction for multiple testing, ** p<0.01. D. The scatter plot shows the correlation of cord IgG1 titer against Spike versus days from maternal booster to delivery. R value reflect a spearman correlation. One dyad was excluded because of missing information about time from booster to delivery.

Ancestral Spike-specific IgG1 levels in the cord were positively correlated with time elapsed since receipt of the booster dose (R=0.57, p = 0.035, Figure 4D). Omicron Spike-specific IgG1 cord levels were also significantly positively correlated with time elapsed since boosting (R=0.68, p = 0.009, Figure S2D).

## Comment

### Principal Findings

Here we show that receipt of a mRNA booster dose induces equivalent IgG1, IgA, and IgM responses to ancestral and Omicron Spike in pregnant, lactating, and nonpregnant individuals. For women who received primary vaccination during pregnancy, a booster dose given in the third trimester significantly increased IgG1 against Omicron. Multivariate analysis revealed differences in Spike-specific epitope coverage and antibody class switching unique to pregnancy. Third trimester boosting resulted in equivalent maternal and cord IgG1 levels, with most efficient transfer of FcγR3a-binding IgG1 to the cord. In individuals boosted in the third trimester, cord IgG1 levels against ancestral Spike and Omicron were positively correlated with increasing time from boost to delivery. Taken together, these results suggest that boosting in pregnancy has the potential to augment SARS-CoV-2 immune protection in pregnant individuals and their neonates, particularly against Omicron.

### Results in Context

Although previously regarded as a generalized immune tolerant state, pregnancy is in fact marked by immunomodulatory changes aimed at balancing wound healing and pathogen surveillance, with tolerance of the fetal allograft.^21,22^ Previous work from our group has demonstrated subtle alterations in the immune response to COVID-19 vaccines during pregnancy compared to nonpregnant individuals as well as differences by trimester of vaccination.^15,23^ The primary differences observed in the immune response between pregnant and nonpregnant individuals after boosting were related to differences in the breadth of epitopes recognized as well as isotype and subclass levels. Specifically, nonpregnant women exhibited a broader targeting of the NTD and S2 domains by functional antibodies, both of which may play a critical role in the general immune response to the Spike antigen. These antibody profiles may confer enhanced immunity in the event of significant mutation in the RBD by VOCs,^24^ and non-neutralizing protection against disease should transmission occur.^25^ Whether additional boosters, or alternative vaccine platforms, might drive enhanced epitope coverage in pregnancy and breach the immunodominance of RBD will be important to assess. The observed enhancement in immunity to RBD, however, is likely key to the augmented protection against VOCs induced by boosting during pregnancy.

Additionally, the significant differences in antibody subclass/isotype selection across nonpregnant and pregnant women in response to boosting may point to a difference in booster-induced B cell selection between pregnant and nonpregnant individuals. Specifically, B cell class switching progresses from IgM to IgG3 > IgG1 > IgA > IgG2 > IgG4.^26^ The selective induction of more functional IgG1 and IgG2 in nonpregnant women points to enhanced functionalization or class-switching in memory IgG B cells. Conversely, the selection of largely IgM/IgG3 responses in pregnant women points to either 1) the selective induction of naïve (new) B cell responses in pregnancy and/or 2) a blockade of further IgG class switching and the interruption of germinal center memory B cell activation and expansion. Given that IgM and IgG3 have potent anti-microbial and anti-viral activity due to their inherent higher affinity for complement and FcRs, respectively,^27,28^ these data may point to a unique hallmark of the immune state that emerges during pregnancy. Interestingly, our data suggest that lactacting women exhibit an intermediate profile between pregnant and nonpregnant women.

Boosting in the third trimester of pregnancy resulted in 1:1 transplacental transfer of total Spike-specific IgG1 to the cord, as evidenced by equivalent levels in the maternal and cord blood at delivery. Although maximal transfer efficiency was not observed for Spike-specific IgG1, i.e. higher antibody concentrations in cord relative to maternal blood,^23,29,30^ this is likely due in part to the relatively short interval between boost and delivery (2-8 weeks). We did, however, observe the selective, efficient transfer of FcγR3a-binding antibodies, which are able to activate natural killer cells, the most mature and functional innate immune cell subset present in the neonate at birth.^31^ Recent data on boosting in pregnancy suggests that compared to natural infection or primary mRNA vaccination in the third trimester, receipt of a booster dose can result in greater maternal and cord IgG titers against the ancestral SARS-CoV-2 Spike at delivery.^32^ Expanding on these data, our results demonstrate that for pregnant individuals vaccinated prior to pregnancy or in early pregnancy, boosting augments IgG1 against ancestral Spike and Omicron, and for those boosted in the third trimester, boosting drives the preferential transfer of highly functional FcγR3a-binding IgG1 to the cord, with greater cord antibody levels observed with increasing time from booster dose to delivery.

### Clinical Implications

The data reported here demonstrating comparable overall immunogenicity of the booster dose in pregnant/lactating compared to nonpregnant individuals may help inform uptake of boosters among these high risk groups, which remain vulnerable to infection with emerging VOCs that can escape neutralization.

Beyond the impact of vaccination on driving pathogen-specific immunity to protect pregnant individuals, vaccine-induced antibody transfer via the placenta is a critical means to provide immunity to the infant.^35,36^ Receipt of a primary COVID-19 mRNA vaccine series during pregnancy is associated with a reduction in newborn hospitalization from COVID-19 in the first 6 months of life,^37^ related to persistent maternal IgG in the newborn circulation.^38^ In this study, pregnant individuals who received a booster dose demonstrated an increase in IgG1 against Omicron compared to levels generated by primary vaccination, and preferential transfer of NK-cell activating FcγR3a-binding antibodies to the cord. In the subset of maternal-neonatal dyads analyzed, in which a booster dose was received in the third trimester, highest cord IgG1 was observed with increased time from boost to delivery. Because COVID-19 vaccines are only recommended for individuals 5 years and older, and infants under 6 months of age will likely not receive a COVID-19 vaccination in the near future, strategies that augment maternal vaccine-induced antibody transfer and optimize infant protection against emerging VOCs are critically important.

### Research Implications

These data pointing to subtle alterations in epitope targeting and subclass/isotype selection in pregnancy suggest that future studies investigating both humoral and cellular immune responses to additional boosters or novel vaccines will be critical to gaining a comprehensive understanding of how the immune response to vaccination is altered in pregnancy. Future investigation into the impact of boosting across all trimesters on the maternal immune response and antibody transfer efficiency at delivery will be important, as will assessment of heterologous boost (receipt of a booster that does not match the primary vaccine received) versus homologous boost in pregnant compared to nonpregnant individuals.

### Strengths and Limitations

Leveraging the systems serology approach, we were able to comprehensively profile the immune response to COVID-19 mRNA boosting across pregnant, lactating, and nonpregnant age-matched controls, enabling both broad and deep assessment of the humoral response to boosting against both ancestral Spike and Omicron. Data on boosting in pregnancy, particularly on the specificity of the antibody response to Omicron, remain extremely limited.

There are several limitations. An approximately 10-day difference in time from booster dose administration to sample collection in nonpregnant women was observed, but is unlikely to be meaningful, as all samples were collected at least two weeks the booster dose was received, when peak responses are expected to occur. All pregnant individuals who delivered during the study period received the booster dose in the third trimester, precluding comparisons of transplacental transfer efficiency by trimester of boosting. Most pregnant individuals in this study were vaccinated prior to conception, limiting our ability to compare the booster response between individuals vaccinated during or before pregnancy. However, the data in our study are most applicable to booster-eligible individuals who are currently pregnant or considering pregnancy in the U.S., as nearly 70% of reproductive-aged women have completed primary vaccination.^39^

## Conclusions

These data suggest that COVID-19 boosting during pregnancy and lactation induces a robust humoral immune response against ancestral and Omicron Spike, comparable to that observed in nonpregnant individuals. Moreover, antibodies were transferred to the cord in a time-dependent manner, suggesting that earlier boosting may be beneficial both for augmenting immunity in the pregnant individual and for optimal transfer of immunity to the infant.

## Data Availability

All data in the present study are available upon reasonable request to the authors.

## Supplementary Figures

**Figure S1.**
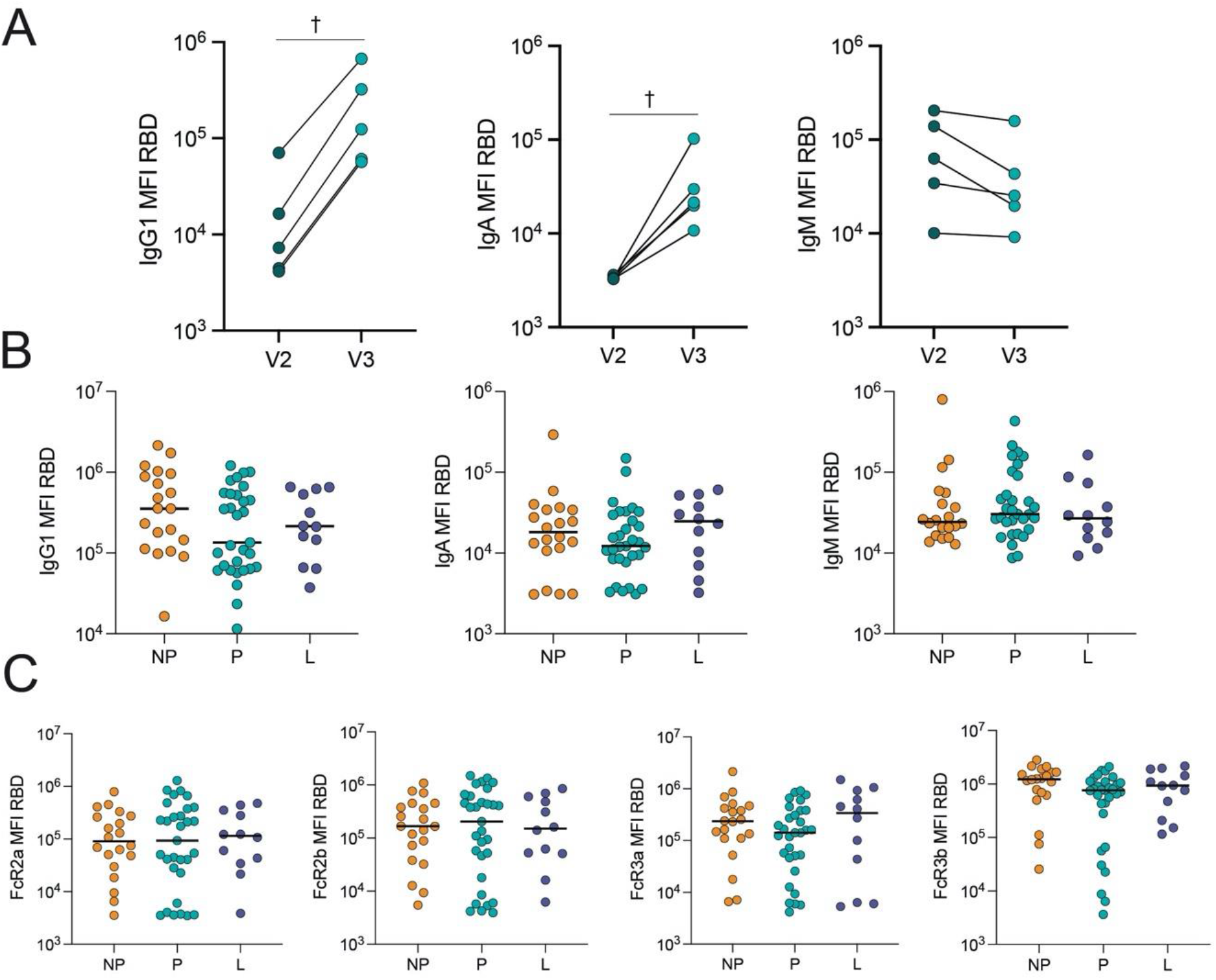
COVID-19 booster vaccine induces a similar Spike-specific RBD antibody response in pregnant, lactating and nonpregnant individuals. A. The dot plots show the peak IgG1, IgA and IgM response against RBD in 5 pregnant individuals after receiving the second dose of a primary mRNA vaccine series (V2) and after the booster dose (V3). Lines connect samples from the same individual. Significance was determined by a Wilcoxon signed-rank test. Differences did not reach statistical significance. † p = 0.06. B. The dot plots show the IgG1, IgA and IgM-titer against RBD in nonpregnant (NP), pregnant (P) and lactating (L) individuals. Horizontal line represents the median for each group. Significance was determined by a Kruskal-Wallis test. No comparisons were significant. C. The dot plots show the FcR-binding of antibodies against RBD in nonpregnant (NP), pregnant (P) and lactating (L) individuals. Horizontal line represents the median for each group. Significance was determined by a Kruskal-Wallis test. No comparisons were significant.

**Figure S2.**
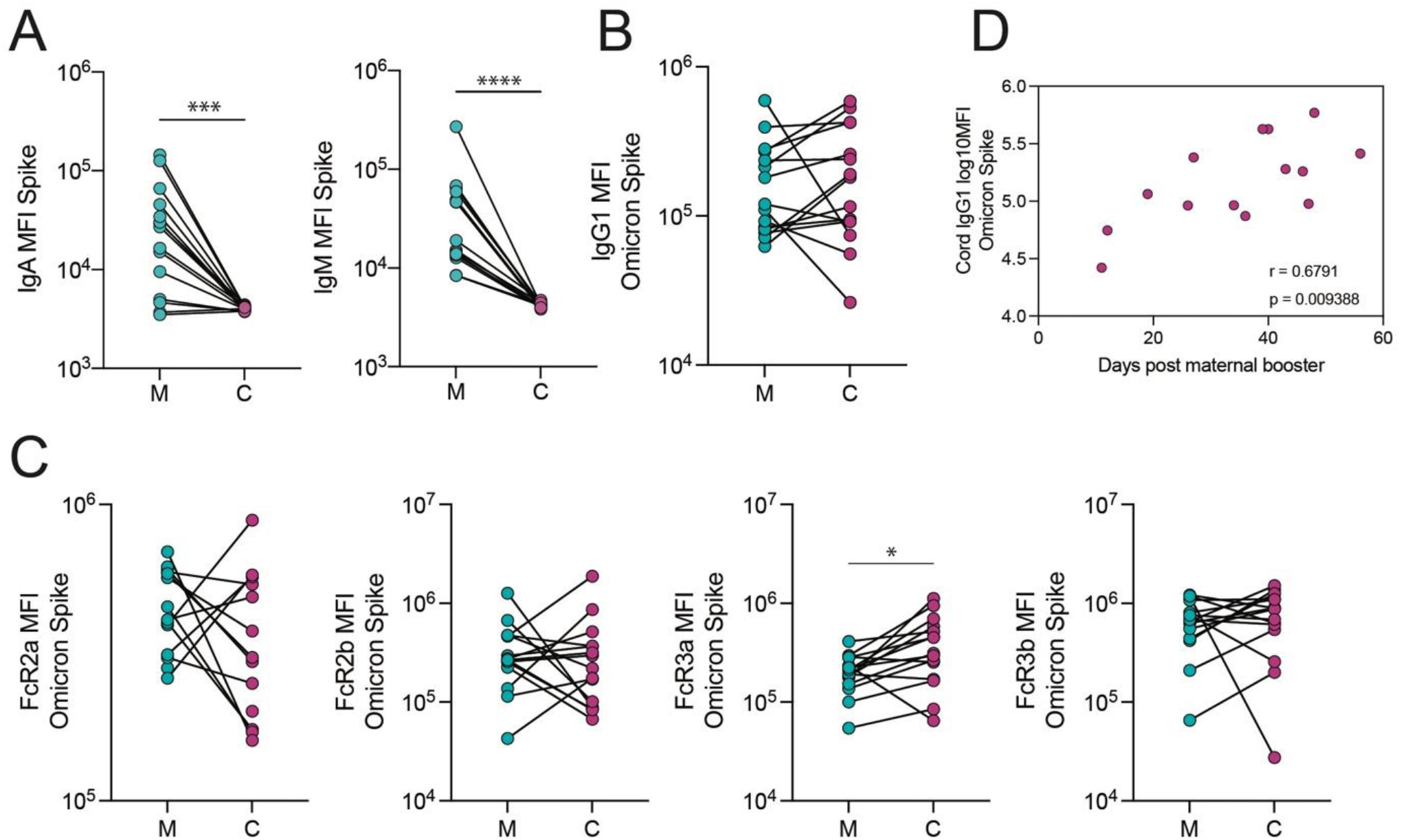
The transfer of Omicron Spike-specific antibodies to the cord after vaccination in the third trimester. A. The dot plots show IgA and IgM against Spike in maternal (M) and cord blood (C). Lines connect matched maternal:cord dyads (n=15). ***p<0.001, ****p<0.0001. B-C. The dot plots show the IgG1 (B) and FcR-binding (C) titer against Omicron Spike in maternal (M) and cord blood (C). Lines connect matched maternal:cord dyads (n=15). Significance was determined by a Wilcoxon signed-rank test followed by a Benjamini-Hochberg correction for multiple testing, * p<0.05. D. The scatter plot shows the correlation of log-transformed cord IgG1 levels against Omicron Spike versus days from maternal booster to delivery. R value reflects a Spearman correlation.

